# Association between sexual violence and depression is mediated by perceived social support among female University students in the Kingdom of Eswatini

**DOI:** 10.1101/2022.09.19.22280102

**Authors:** Rebecca Fielding-Miller, Lotus McDougal, Elizabeth Frost, Sakhile Masuku, Fortunate Shabalala

## Abstract

**Background:** Gender-based violence is a tool that primarily functions to maintain gendered power hierarchies. As manifestations of gender-based violence, sexual assault and street harassment have been shown to have significant effects on mental wellbeing in the global North, however there is little research centering the experiences and consequences of gendered harassment in the Africa region.

**Methods:** We analyzed a cross-sectional random sample of women attending a major university in Eswatini in 2017 to measure the prevalence of street harassment among female university students and assess the relationship between experiences of sexual assault, sexualized street harassment, and mental health outcomes in this population.

**Results:** We found that in the previous 12 months, women reported experiencing high levels of sexual assault (20%), street harassment (90%), and depression/anxiety (38%). Lifetime sexual assault, past 12 months sexual assault, and street harassment were all significantly associated with symptoms of depression. We created a structural model to test hypothesized causal pathways between street harassment, previous experiences of sexual assault, and symptoms of depression, with social support as a potential mediator. We found that a history of sexual violence significantly mediated the association between street harassment and depression, and that social support mediated a large proportion of the association between both forms of gender-based violence and depression.

**Conclusion:** Sexualized street harassment is associated with increased anxiety and depression for nearly all women, however the effects are especially pronounced for women who have previous experiences of sexual violence. Sexualized street harassment functions as a tool to maintain gendered power hierarchies by reminding women of ongoing threat of sexual violence even in public spaces. Social support and solidarity among women is a potentially important source of resiliency against the physical and mental harms of all forms of gender based violence.

## Background

Gender-based violence encompasses a spectrum of behaviors which all function to reinforce gendered power hierarchies. Sexual assault, harassment, and street-based verbal harassment all fall within this spectrum (1). The public nature of street harassment reinforces gendered power structures by providing perpetrators the opportunity to perform sexually aggressive behaviors congruent with toxic hegemonic masculinity while simultaneously reinforcing women’s status as sexual objects who are vulnerable to control by sexual violence and potentially unsafe in public spaces (2, 3). Experiencing street harassment has been associated with increased depression, anxiety, inability to sleep, disempowerment, humiliation and fear of public spaces (4-9).

A growing body of literature is emerging on gender-based violence among university women both on and off campus(10-14). Social movements such as #MeToo have increased awareness of the prevalence of gender-based violence across many contexts, with women across the globe coming forward to speak openly about their experiences. Likewise, in many high income countries we have seen improved reporting mechanisms become available for students who fall victim to sexual assault or sexual harassment, and university policies that have been reconstructed with a lower tolerance for instances of sexual violence (15) However, much work is still needed to curb the high rates of behaviors across the spectrum of sexual violence for women attending university, particularly in settings such as Southern Africa where rates of sexual violence remain high (16-18).

Sexual harassment is both a manifestation of sexual violence and a tacit threat of future sexual assault, and as such, can have serious effects on women’s willingness to participate in public spaces. When this form of gender-based violence manifests in secondary and higher educational settings, the potential for adverse downstream consequences are serious. Sexual harassment in academic settings can negatively affect academic performance, which in turn reduces women’s future workforce participation, compromising future individual earnings as well as social benefits (19, 20). Women may feel unsafe or unwelcome as they travel to school (21), engage in coursework (22), or pursue mentorship or higher level training (19, 20, 23).

Previous studies have established that a history of sexual assault corresponds with higher risk of depression, anxiety and post-traumatic stress disorder (24-26). As with sexual harassment, a history of sexual assault can also compromise academic attendance and performance, as well as subsequent relationships (27). Sexual assault before the age of 18 years has been linked to increased levels of heavy alcohol use, suicide ideation and suicide attempts(28, 29).

Efforts to operationalize and measure the prevalence of street harassment are relatively new, however existing reports suggest that street harassment is extremely prevalent (9, 30, 31). Data on street harassment from low and middle income countries (LMICs) are less robust: a 2021 systematic review and meta-analysis of sexual harassment in LMICs found that sexual harassment was both extremely common and associated with higher rates of anxiety and depression. Relatively few of these studies focused on harassment in public, however. We know of no studies assessing the prevalence and statistical associations between poor mental health and street harassment in southern Africa.

While each woman’s experience of sexual violence and its aftermath is unique, the relationships between GBV (including harassment) and poor mental health outcomes are influenced by factors at individual, relational, and structural levels (32, 33). A robust body of evidence demonstrates that social support plays a particularly important role in mediating the association between sexual violence and adverse mental health outcomes (34), although there is less research exploring how social support might potentially influence the link between sexual or street harassment and mental health.

The goal of the present study was to assess the effects of sexual assault and street harassment on mental health among female university students in Eswatini and determine the potential role of social support as a mediating factor.

## Methods

### Study Setting, Sample and Design

Eswatini is a small Kingdom in Southern Africa with a population close to 1.1 million (35). In Eswatini, one third of girls have experienced sexual abuse by the age of 18 years and close to half of women experience gender-based violence in their lifetime (36). The current study is part of a larger project to measure the prevalence and correlates of campus sexual assault at the University of Eswatini (UNESWA)(37). UNESWA enrolls approximately 7,500 students, most of whom are undergraduates. The main campus, Kwaluseni, enrolls approximately 4500 students, 30% of whom live on campus with the remainder in peri-urban areas nearby. Women make up 58% of the total student population but are 15% more likely to drop out before their 4th year than men64. The majority of students are Swazi citizens. However, the university enrolls students from throughout the region including South Africa, Botswana, and Zimbabwe, and the language of instruction is English. Women were eligible to participate if they spoke English, were enrolled full time, and were at least 18 years old. Selected participants were contacted via email, text, and phone calls inviting them to visit the on-campus student health clinic to complete the consent process. Participants were randomly selected from the University Register’s list of all full-time female students enrolled at the primary UNESWA campus. After indicating that they understood the risks and benefits of the study, participants were asked to complete a behavioral survey using a tablet with computer-assisted self-interview (CASI) software. Participants completed the survey in a private office, with a trained research assistant nearby to provide assistance and answer questions. Data collection occurred from February – November 2018.

### Ethics

The study was reviewed and approved by UNESWA Research Ethics Board and the University of California, San Diego Institutional Review Board (IRB). Data collection procedures followed the World Health Organization’s Recommendations for Intervention Research on Violence Against Women. All participants were offered a 25 Emalangeni (approximately $2 USD) honorarium to thank them for their time and expertise. A research assistant who was trained in trauma informed counselling was always available while participants completed the survey, and all participants were given a card with a phone number for free-of-charge tele-counselling services when they left the study site. The study team made pro-active efforts to reduce vicarious trauma in frontline staff, including engaging in regular debriefing sessions and ensuring that a local, no-cost third party counselor was available to all staff members.

### Measures

Our primary outcome was depression, assessed using the 10-item Center for Epidemiological Studies Depression Scale (CES-D-10). The CES-D-10 asks participants to report on their experiences and symptoms in the previous seven days and has been validated in the southern African context (38). The scale ranges from possible scores of 0-30, and previous validation work in the region suggests that participants are likely experiencing depression or anxiety if they score 12 or above (38).

Our primary predictors of interest were women’s self-reported experiences of (1) sexual assault and (2) street harassment. We measured sexual assault using the sexual experiences survey short form (SES-SF) (39), a behavioral scale that asks participants to report on their experiences of completed and attempted sexual assault in their lifetime and past 12-months. Women were categorized as having experienced sexual assault if they reported non-consensual penetration by means of threats, coercion, physical violence, or being incapacitated. Street harassment was measured by asking women “how often do you experience unwanted comments from men in public” with the options of “never”, “less than once a month”, “once a month”, a few times a week”, and “every day”.

Social support was assessed using three survey items, each with a 4-point Likert response: “If I were sexually assaulted by a [stranger / acquaintance / boyfriend or person I have spent time alone with] I have friends who would support me.” Women could respond “definitely yes,” “probably yes,” “probably no,” and “definitely no.”

### Analyses

We conducted univariate descriptive analyses for our primary outcome (CES-D-10 scale) and hypothesized predictors (lifetime penetrative sexual assault, street harassment, perceived social support), followed by bivariate analysis of variance (ANOVA) tests to assess the difference in mean CES-D-10 scores across predictors.

We then used STATA 16.0 software(40) to build two confirmatory factor analysis (CFA) measurement models assessing (1) the validity of the CES-D-10 as a latent construct in the population of interest, and (2) the three social support items as a coherent construct of perceived social support following a sexual assault. Indicators that loaded with a value below 0.50 in these CFA models were removed, and measurement error covariances were added based on theoretical considerations and after considering modification indices suggested by the *estat mindices* command. *Estat mindices* uses Lagrange multiplier tests to suggest additional covariances or path coefficeints that would significantly improve model fit(41). With each modification to the CFA measurement model, we used chi-square tests to assess whether the nested model had a statistically significant improved fit to the data compared to the previous iteration at p <0.05. After finalizing the measurement models we constructed a full structural equation model to test the hypotheses that previous experiences of sexual assault and perceived social support significantly mediated the association between experiences of sexualized street harassment and depression.

We used a full-information maximum likelihood (FIML) estimation strategy (*mlmv* estimation command) for both the measurement and full structural equation models to account for the presence of ordinal indicators in the depression and social support latent variables. Overall model goodness of fit for measurement and the full structural model was assessed across multiple fit indices, per Kline’s recommendation: Model chi-square, RMSEA and RMSEA 90% confidence interval, and the Bentler Comparative Fit Index (CFI)(42). Direct, indirect, and total effects were examined using Stata’s *teffects* command.

## Results

Three-hundred and seventy-two women participated in the survey (response rate of 49.5%). Mean participant age was 23.3. years, (range 18-45, standard deviation: 3.7). Three hundred and eighteen participants completed all 10 CES-D-10 items, with a median score of 9.5 (range: 0-30, SD: 5.6). There were no significant differences in outcomes of interest between participants who did (n=318) and did not (n= 54) complete all CES-D-10 items (analyses not shown). Thirty-one percent of participants (n=97) who completed all CESD-10 items reported experiencing penetrative sexual assault in their lifetime and the vast majority (90%) reported experiencing unwanted sexual comments from men in public in the previous year. The majority of study participants felt that they had a friend who would ‘definitely’ support them if they were assaulted by a stranger (62%), acquaintance (60%), or boyfriend (54%).

There were statistically significant associations between all hypothesized predictors and reported depressive symptomology. The median CES-D score for women who reported ever experiencing penetrative sexual assault was 12.0 compared to 9.5 for women who never experienced penetrative assault (p<0.01). There was also a significant dose response relationship between the frequency of unwanted sexual comments from men in public and CES-D scores, ranging from a median CES-D score of 8.1 for women who reported ‘never’ receiving comments, to 14.0 for women who reported receiving comments ‘every day’ (p<0.001). There were similar linear associations between depressive symptoms and perceived social support in the instance of assault from a stranger, acquaintance, and boyfriend or boy with whom the respondent had spent time alone (all p<0.001) (Table 1).

**TABLE 1:**
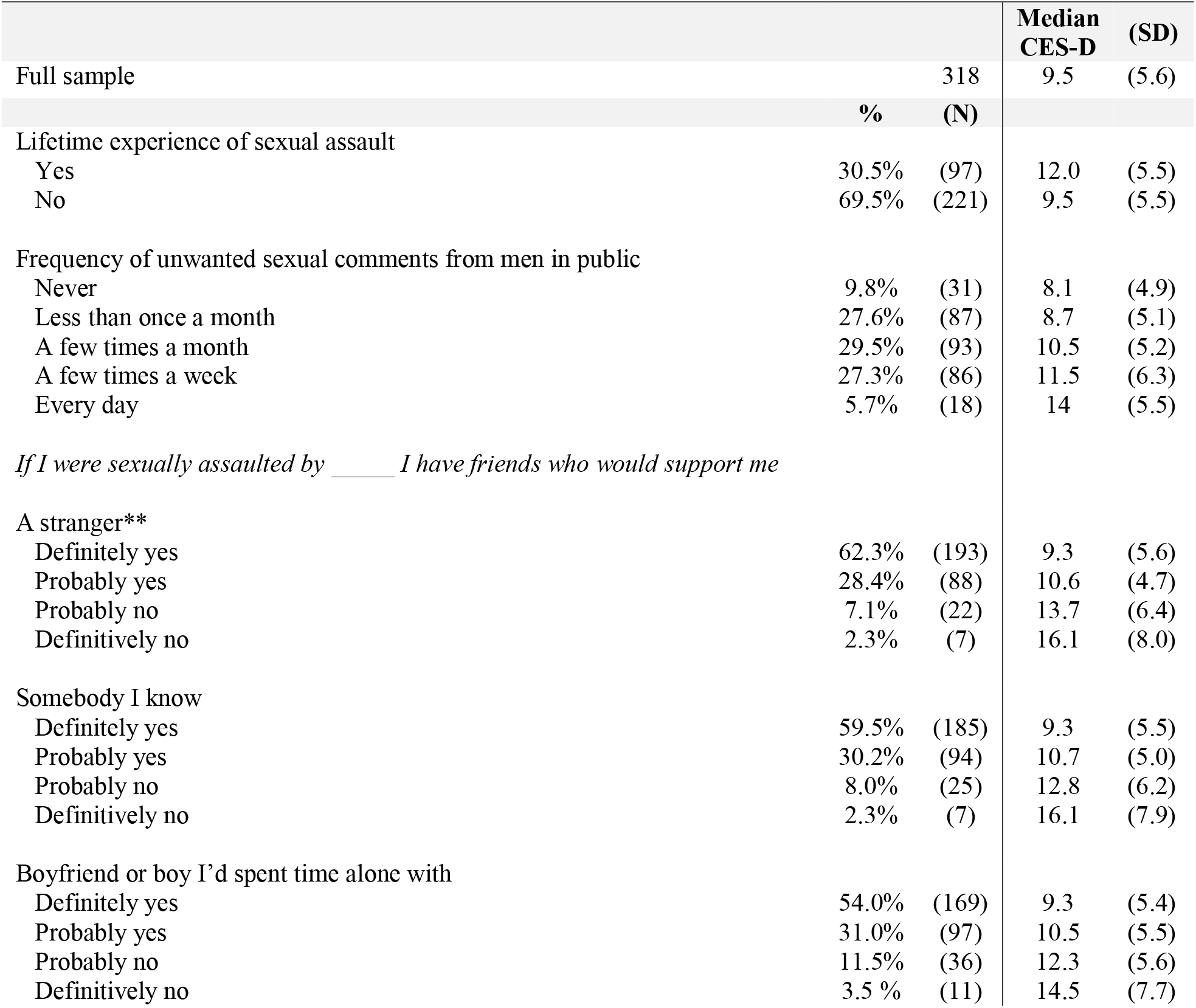
Lifetime sexual assault, street harassment, and perceived social support by median Center for Epidemiological Studies Depression (CES-d) score.

We next constructed measurement models to assess how well all 10 CES-D items fit our data as a single latent construct of depression. Two of the ten indicators (“everything I did was an effort” and “I felt hopeful about the future.”) had a factor loading below 0.50. After removing these two indicators, the more parsimonious model fit the data approximately as well as the 10-item model (chi-square = 40.0, df = 20 vs. chi-square = 103.6, df = 35), and so we chose to measure the latent depression construct using the more parsimonious 8 item model. We further modified the model to correlate error measurements between “I was bothered by things that don’t usually bother me” and “I had trouble keeping my mind on things I was doing,” as well as “I felt depressed” and “I had trouble sleeping” and then “I felt lonely” and “I could not get going.” (Figure 1a). The final CFA measurement model fit the data well, with all factor coefficients greater than 0.50 (p<0.05), chi-square = 18.6 (df = 17, p = 0.60) RMSEA = 0.02, and CFI = 0.998. The social support measurement model was just-identified, with 0 degrees of freedom. All 3 social support indicators loaded at 0.80 or above, with p<0.001 (Figure 1b)

**Figure 1a:**
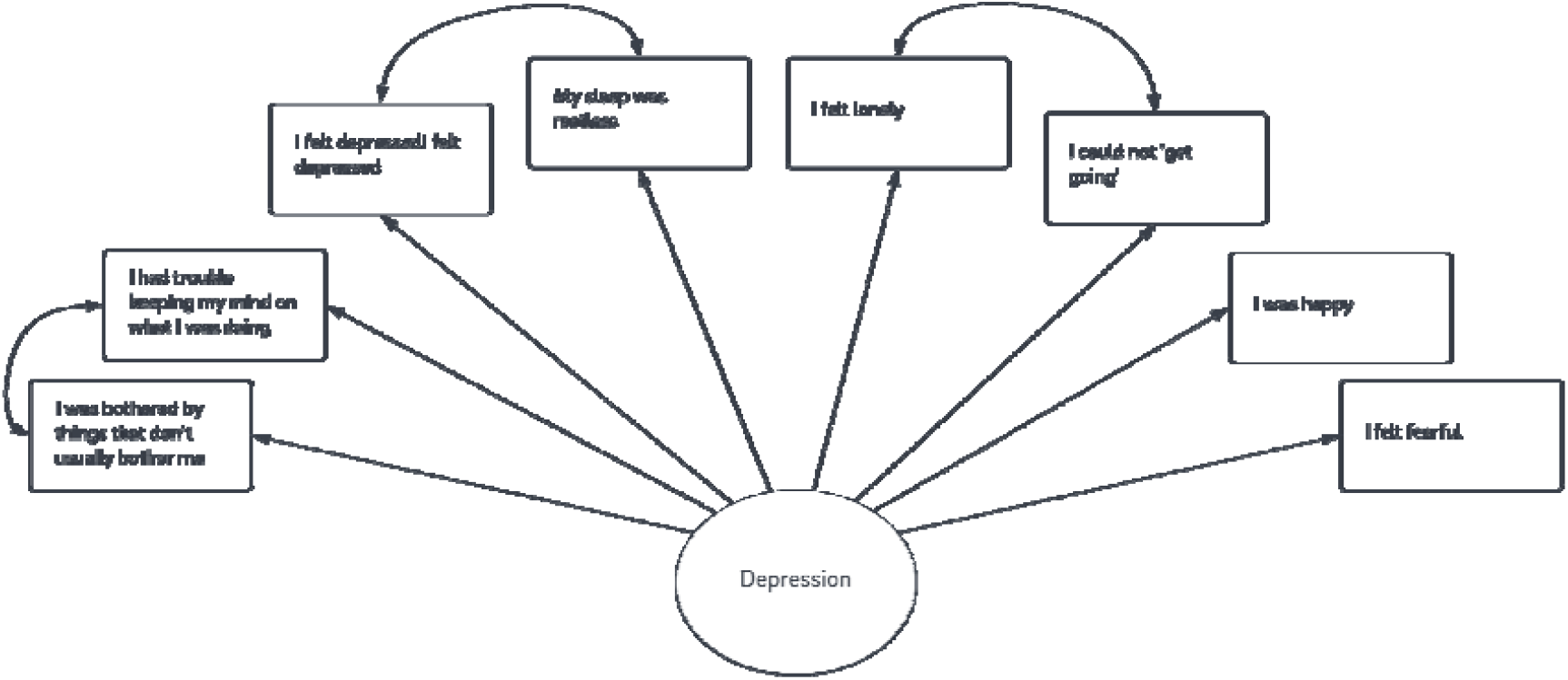
Measurement model of depression as a latent construct

**Figure 1b:**
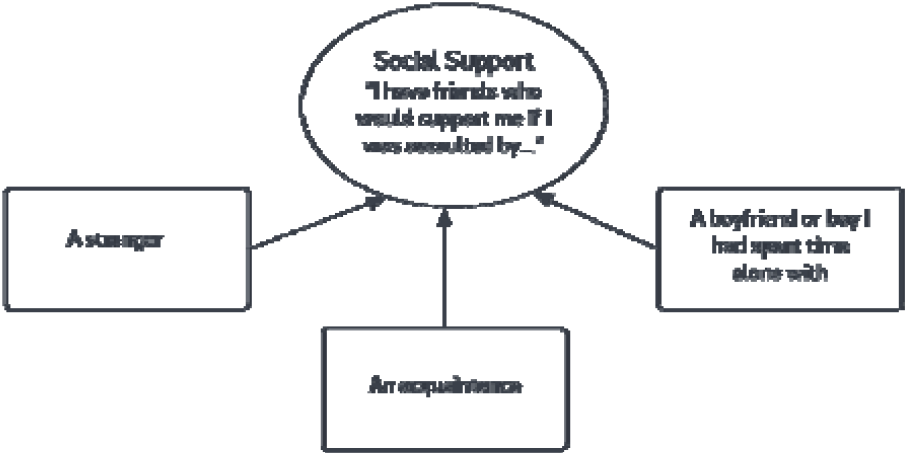
Measurement model for social support as a latent construct

The fully specified structural model fit the data well, with chi-square = 74.4, (df = 58, p = 0.07), RMSEA = 0.03 (90% CI: 0.00 – 0.05), and CFI = 0.99 (Figure 2). Confidence that a friend would support her in the case of sexual assault had the largest direct effect on the probability of experiencing depression. For each standard deviation increase in perceived social support, the probability of experiencing depression decreased by 0.27 standard deviations (standardized *b* = −0.27, 95% CI: −0.37 -- −0.17). Experience of lifetime penetrative sexual assault was associated with a 0.11 standard deviation increase in the probability of depression (standardized *b* = 0.11, 95% CI: 0.01 – 0.22) and one standard deviation increase in the reported frequency of sexualized street harassment was associated with a 0.19 standard deviation increase in the probability of depression, (standardized *b* = 0.19, 95% CI: 0.09 – 0.30). Women who reported experiencing street harassment were significantly more likely to also report a lifetime experience of penetrative sexual assault (standardized *b* = 0.13, 95% CI: 0.03 – 0.23), and were less certain that a friend would support them after an experience of assault (standardized *b* = 0.28, 95% CI: 0.18 – 0.38) (Table 2).

**TABLE 2:**
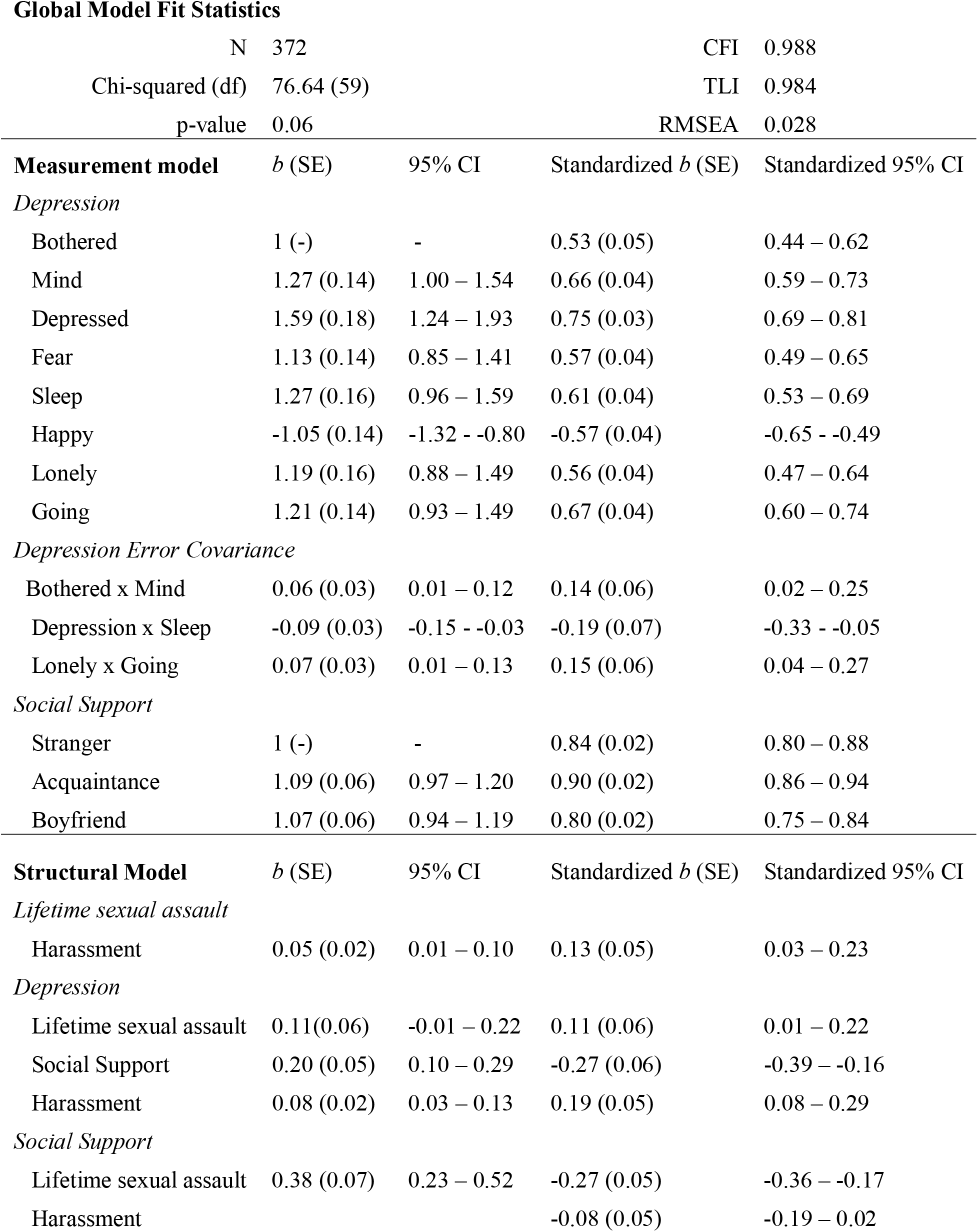
Model parameters and fit statistics

**Figure 2:**
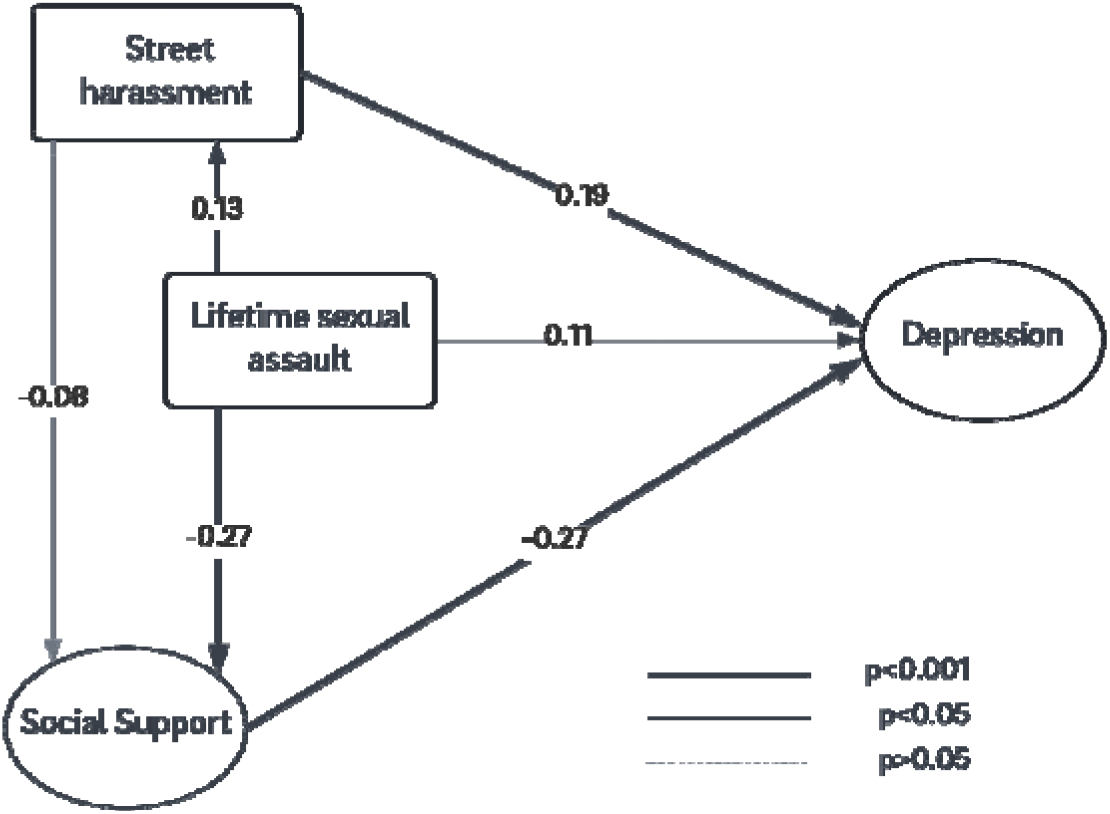
Structural Equation Model with standardized path coefficient

Decomposed direct and indirect standardized effects of each covariate are shown in Table 3. When we examined indirect effects, we found that the social environment played a significant mediating role in the association between sexual assault and depression. Thirty-nine percent of the association between lifetime experience of sexual assault and depression in the previous two weeks was mediated by perceived social support, while 22% of the association between experiencing sexualized street harassment and depression was mediated by having a lifetime experience of sexual assault.

**TABLE 3:** Structural model pathway covariates and proportion of pathway mediated

|  | Direct *b* | p-value | Indirect *b* | p-value | Total *b* | p-value | Proportion Mediated |
| --- | --- | --- | --- | --- | --- | --- | --- |
| **Depression** |  |  |  |  |  |  |  |
| Sexual assault | 0.11 | (0.05) | 0.07 | (0.001) | 0.18 | (0.001) | 0.39 |
| Social Support | -0.27 | (<0.001) | --- | --- | -0.27 | (<0.001) | --- |
| Harassment | 0.19 | (0.001) | 0.05 | (0.02) | 0.23 | (<0.001) | 0.22 |
| **Social Support** |  |  |  |  |  |  |  |
| Sexual assault | -0.27 | (<0.001) | --- | --- | -0.27 | (<0.001) | --- |
| Harassment | -0.08 | (0.14) | -0.03 | (0.03) | -0.11 | (0.04) | 0.27 |
| **Sexual Assault** |  |  |  |  |  |  |  |
| Harassment | 0.13 | (0.01) | --- | --- | 0.13 | (0.012) |  |

## Discussion

Depression, sexual assault, and frequent sexual harassment were pervasive in this sample of Swazi female undergraduates. Over 38% reported symptoms consistent with depression in the previous week, with the prevalence significantly higher amongst those who had experienced sexual assault in their lifetime, and amongst those who reported higher rates of street-based sexual harassment. Close to 90% of women reported experiencing unwanted sexual comments in public, with nearly 30% experiencing this harassment on a weekly basis. This prevalence is comparable with data reported from the United States which used a similar measure of street harassment (43). Our findings suggest that the social environment in which women live, work, and attain their education can have a significant effect on the mental health impact of sexual assault. A primary function of sexualized street harassment is to reinforce patriarchal dominance via an ever-present threat of sexualized male violence and humiliation. This experience of harassment is likely to be especially salient for women who have previous experiences of sexual assault. In contrast, a supportive social environment may serve as a key aspect of resilience, significantly mediating the pathway between sexual violence and depressive symptoms.

Social support has been shown to mitigate the adverse mental health consequences of sexual harassment and assault in other settings (44, 45), underscoring its potential as a strategy for collective resistance against sexual violence as a tool of gendered power and control (46). Notably, while approximately 90% of women reported that they had friends who would ‘definitely’ or ‘probably’ support them if they disclosed assault by stranger or an acquaintance, the number was slightly lower in the case of assault by a boyfriend (85%). While this difference was not statistically significant, marital rape was only outlawed in the Kingdom of Eswatini in 2019, and university women have previously reported the difficulty associated with disclosing assaults committed by an intimate partner (47). This is consistent with the ways in which Connell has articulated cathexis, or the role of emotional and affective relationships in maintain gendered power structures (48). While a pre-existing romantic, sexual, or otherwise social relationship obviously does not negate the possibility of sexual assault (and is indeed the most common setting for sexual violence), it may dampen women’s access to social support and willingness to disclose an event.

Women who were more certain that their friends would support them in the event of a sexual assault reported lower levels depressive symptoms in the previous two weeks. It is important to note that this association is a correlation within a cross-sectional study and cannot be interpreted as causal. Low levels of social support have been found to be predictive of repeat sexual assault in some university populations, (49) suggesting that women with lower levels of social support may be more vulnerable to additional experiences of gender-based violence. Moreover, the association between sexual violence and perceived sexual support may be a result of lived experience – it is possible that women who have not been sexually assaulted may be more optimistic about the likelihood that their friends will support them, while women who have been assaulted may be more likely to report that they did not, in fact, receive social support when it was needed, or may fear friends’ reaction if they do disclose.

The findings of this study should be interpreted within the study’s limitations. Our response rate, at 49.5%, was low for a demographic survey, although it is quite high for a campus sexual assault survey (50). Survey data are self-reported, and therefore vulnerable to both recall and social desirability bias. However, both of these biases were mitigated to the extent possible by asking about current or past year experiences, and by using self-administered surveys in a private office (51).

To our knowledge, this is the first documentation of the association between verbal sexual street harassment and adverse mental health amongst young women in the southern African context, and is in line with findings elsewhere as well as social movements such as #MeToo (4, 5, 43, 52-56). The ubiquity of street harassment faced by women is a reminder that women often lack the autonomy and normative right to safely occupy public spaces. Research on male perpetrators of street harassment indicates that they harass in order to control women, and that they are more likely to harass when they feel they have less power than women (3). Our data lends additional credence to the notion that sexualized street harassment exists as part of a spectrum of sexual violence, the function of which is primarily to reinforce gendered social control. Bystander interventions could play an important role in addressing the social norms surrounding street harassment. Unfortunately, to date very few (if any) interventions exist which specifically aim to prevent perpetrators from engaging in street harassment, or to support women and/or gender non-conforming individuals to engage in resistance strategies when they experience harassment (1). More focused intervention and support strategies have the potential to meaningfully improve women’s safety, bodily integrity, and mental health.

## Data Availability

Data are available at https://dataverse.harvard.edu/dataset.xhtml?persistentId=doi:10.7910/DVN/WJPGIL

## Declarations

### Ethics approval and consent to participate

All research was performed in accordance with the Declaration of Helsinki. The study was reviewed and approved by UNESWA Research Ethics Board and the University of California, San Diego Institutional Review Board (IRB) (Federalwide Assurance 00004495; protocol 181603). Data collection procedures followed the World Health Organization’s Recommendations for Intervention Research on Violence Against Women. All participants provided informed consent previous to completing a survey. A research assistant who was trained in trauma informed counselling was always available while participants completed the survey. The study team made pro-active efforts to reduce vicarious trauma in frontline staff, including engaging in regular debriefing sessions and ensuring that a local, no-cost third party counselor was available to all staff members.

### Consent for publication

NA

### Availability of data and materials

The datasets used and/or analysed during the current study are available from the corresponding author on reasonable request.

### Authors contributions

RFM: Conceptualization, methodology, formal analysis, data curation, writing (original draft), supervision, funding acquisition; LM and EF: Writing (review and editing); SM: Conceptualization, project administration, writing (reviewing and editing); FS: Conceptualization, methodology, resources, writing (reviewing and editing), supervision, funding acquisition.

## Acknowledgments

The study team wishes to thank the women who shared their time and stories with us to make this research happen. We are indebted to their generosity of spirit. We thank the staff and administration of the University of Eswatini for their willingness to support this work and desire to build an evidence-based response to sexual violence on campus. RFM wishes to thank her research assistant, Ms. Esther Krohne, who made vital contributions to survey programming and in-person training activities.

